# Olanzapine inhibits hepatic apolipoprotein A5 secretion inducing hypertriglyceridemia in schizophrenia patients and mice

**DOI:** 10.1101/2021.02.26.21252514

**Authors:** Xiansheng Huang, Yiqi Zhang, Wenqiang Zhu, Piaopiao Huang, Jingmei Xiao, Yang Yang, Li Shen, Fei Luo, Wen Dai, Rong Li, Renrong Wu

## Abstract

Olanzapine, an antipsychotic drug, was reported to induce hypertriglyceridemia, whereas the underlying mechanism remains incompletely understood. This study was to determine the role of apolipoprotein A5 (apoA5) in olanzapine-induced hypertriglyceridemia. In this study, 36 drug-naive and first-episode schizophrenic adult patients (aged 18–60 years) in a multi-center clinical trial (ClinicalTrials.gov NCT03451734) were enrolled. Before and after olanzapine treatment, plasma lipid and apoA5 levels were detected. Moreover, 21 female C57BL/6 J mice (8 weeks old) were divided into 3 groups (n = 7/each group): low-dose olanzapine (3 mg/kg/day), high-dose olanzapine (6 mg/kg/day) and control group. After 6 weeks, plasma glucose, lipids and apoA5 as well as hepatic apoA5 protein and mRNA expression in these animals were detected. In our study *in vitro*, primary mouse hepatocytes and HepG2 cells were treated with olanzapine of 25, 50, 100 μmol/L, respectively. After 24 hours, apoA5 protein and mRNA levels in hepatocytes were detected. Our study showed that olanzapine treatment significantly increased plasma triglyceride levels and decreased plasma apoA5 levels in these schizophrenic patients. A significant negative correlation was indicated between plasma triglyceride and apoA5 levels in these patients. Consistently, olanzapine dose-dependently increased plasma triglyceride levels and decreased plasma apoA5 levels in mice. Surprisingly, an elevation of hepatic apoA5 protein levels was detected in mice after olanzapine treatment, with no changes of *APOA5* mRNA expression. Likewise, olanzapine increased apoA5 protein levels in hepatocytes *in vitro*, without changes of hepatocyte *APOA5* mRNA. Therefore, our study provides the first evidence about the role of apoA5 in olanzapine-induced hypertriglyceridemia. Furthermore, plasma apoA5 reduction, resulting in hypertriglyceridemia, could be attributed to olanzapine-induced inhibition of hepatic apoA5 secretion.

## Introduction

Schizophrenia is a severe mental disorder with an approximate lifetime prevalence of 1.0% [1]. Atypical antipsychotics are commonly prescribed as a first-line treatment for schizophrenia because of their effectiveness and much lower rates of extrapyramidal side effects comparing with typical antipsychotics [2]. However, atypical antipsychotics have more metabolic side effects such as weight gain and disturbed glucose and lipid metabolism. Olanzapine is one of the most prescribing atypical antipsychotics for schizophrenia patients [3] and produces overt dyslipidemia that remarkedly increases the morbidity and mortality of coronary heart disease (CHD) [4, 5]. Previous studies have shown that olanzapine-induced dyslipidemia is characterized with elevation of plasma triglycerides, low-density lipoprotein cholesterol (LDL-C) and total cholesterol, as well as reduction of high-density lipoprotein cholesterol (HDL-C) [6,7]. Of note, hypertriglyceridemia dramatically increases the CHD risk for schizophrenia patients [8-10]. The pathogenetic mechanism of olanzapine-induced hypertriglyceridemia is still not elucidated, although it might be associated with central histamine H1 antagonism and increased appetite or with direct impairment of metabolic regulation and alteration of insulin sensitivity.

Apolipoprotein A5 (apoA5), which is a liver-specifically synthesized and secreted protein and considered as a novel member in apolipoprotein family, has been proven to be an important regulator in triglyceride metabolism. Increasing evidences indicate that apoA5 is a potent regulator of triglyceride despite its low concentration in plasma [11-15]. ApoA5 exerts an atheroprotective effect via decreasing plasma levels of triglycerides and triglyceride-rich lipoproteins [16-18]. Fibrates, a widely-used triglyceride-lowering drug, ameliorates hypertriglyceridemia by up-regulating hepatic apoA5 expression [19, 20]. Specifically, *APOA5* genetic polymorphisms are associated with olanzapine-induced dyslipidemia [21, 22]. These data suggest a potential role of apoA5 in olanzapine-related dyslipidemia that was to be investigated in the present study.

## Methods

### Human studies

#### 1. Participants

This study was conducted in The Second Xiangya Hospital Central South University, China from January 2018 to June 2019. Participants were assessed for schizophrenia in accordance with criteria established in the Diagnostic and Statistical Manual of Mental Disorders-Fifth Edition (DSM-5; American Psychiatric Pub; 2013) [23]. Moreover, schizophrenia patients aged 18–60 years in their first psychotic episode of schizophrenia were included in this study.

Exclusion criteria were as following: 1) concurrent diagnosis of other psychiatric disorders defined in DSM-5; 2) planning to be pregnant, pregnancy or breastfeeding; history of alcohol, cigarettes or other substance use; known medical conditions that might affect metabolism; 3) history of diabetes, hypertension, other cardiovascular diseases, endocrine diseases, lipid disorders or serious chronic diseases (such as liver and kidney dysfunction, heart failure, *etc*.). This study was performed in accordance with the Declaration of Helsinki [24], and approved by the Ethics Committee of The Second Xiangya Hospital, Central South University. After a complete description of the study to the participants, informed consent was obtained prior to study participation.

#### 2. Intervention

The participants were assigned to an 8-week treatment with olanzapine (15-20mg/day at 8:00 p.m.). The dose of olanzapine initiated with 5mg/day and then titrated to 15-20mg/day in the first week.

#### 3. Assessments

All patients who received the treatment were scheduled to have a clinical evaluation through the scheduled follow-up at weeks 4 and 8, and 12. The baseline assessments included demographics, a comprehensive medical history, physical examination, anthropometric measurements (weight and height), and PANSS score. The laboratory tests at baseline included fasting lipids and glucose, liver and renal function, blood counts, and electrocardiogram. At each follow-up visit, all baseline clinical evaluations, including physical examination, anthropometric measurements, and laboratory tests, were repeated. The Treatment Emergent Symptom Scale (TESS) [25] was used to record adverse events throughout the clinical trial. PANSS were also evaluated at the end of the trial.

Primary outcomes were the change in fasting plasma triglyceride and apoA5. Levels of plasma apoA5 were determined using ELISA kits from Novus Biologicals (NBP2-68250, USA). The blood glucose and lipids (including triglycerides, cholesterol, LDL-C and HDL-C) were detected by automatic biochemical analyzer.

Secondary outcomes were: 1) the increase in the levels of other lipids, which included HDL-C, LDL-C, total cholesterol; 2) the change of fasting glucose and body mass index (BMI); and 3) psychopathologic symptoms measured by positive and negative symptom scales (PANSS). BMI is calculated as weight in kilograms divided by height in meter squared.

#### 3. Statistical Analyses

All continuous variables with normal distribution were expressed as the mean ± standard error. Changes of parameters (*i*.*e*., body mass index, plasma lipid and apoA5 levels) were defined as the amount of their alterations from baseline to endpoint (*i*.*e*., 4 and 8 weeks). The continuous variables were analyzed with ANOVA. Pearson analysis was used for correlation test. The P value was used to indicate statistical difference, and P < 0.05 was considered statistically significant.

Pearson analysis was used for correlation test. The P value was used to indicate statistical difference, and P < 0.05 was considered statistically significant. Graphpad Prism 8.0.1 statistical software and SPSS(Statistical Product and Service Solutions) were used for statistical analyses of all the data.

### Animal studies

#### 1. Animals

Twenty-one female C57BL/6 J mice (8 weeks old) were purchased from Hunan Stryker Jingda Animal Co., Ltd. All animals were housed under standard conditions with individually ventilated cages with an artificial 12:12-hour light/dark cycle (lights on: 8:00 am) at room temperature (20 − 25°C) and fed *ad libitum* with free access to water. Mice were fed with a regular chow diet containing 4.5% fat (0.02% cholesterol) throughout the experimental period. All experiments were performed in accordance with the National Institute of Health Guide for the Care and Use of Laboratory and approved by the Experimental Animal Ethics Committee of The Second Xiangya Hospital, Central South University.

#### 2. Treatments

Administration of drugs was initiated after one week of acclimatization. The mice were randomly categorized into olanzapine and control group. Mice in the low-dose olanzapine group (n = 7) received 3 mg/kg olanzapine (LY170053, MCE, USA) per day through gavage for 6 weeks, and mice in the high-dose group were treated with 6mg/kg for 6 weeks. Control group was administered with 0.01 ml/g dimethyl sulfoxide (DMSO) once per day by gavage. Olanzapine was dissolved in 0.1% DMSO. The set of olanzapine dosage in this study was based on previous studies [26-31].

#### 3. Determination of body weight, sample collection and biochemical analyses

Body weight of all animals were assessed weekly during the study. At 6 weeks, animals were fasted for 4 hours and then anesthetized by pentobarbital, the liver samples were collected and frozen in liquid nitrogen immediately and then stored in a −80 °C freezer for subsequent analyses. The blood samples were collected from heart in EDTA-coated tubes and then centrifuged at 4 °C at 1000 g for 15 min to isolate the plasma. The plasma levels of glucose, triglycerides and total cholesterol were measured using Spotchem EZ SP 4430 (ARKRAY, Inc., Kyoto, Japan). The plasma apoA5 levels were determined using ELISA kits from Chemical Book (A104763, China).

#### 4. Glucose Tolerance Tests

At the end of the study, all animals underwent a glucose tolerance test (GTT). Mice were fasted for 6 hours with water provided *ad libitum* from 9 am on the experimental day. Initial blood glucose levels of mice were collected 2 hours prior to GTT. During GTT, blood glucose levels were obtained at 15, 30, 60, 90, and 120 minutes after an *i*.*p*. dose of glucose (1.5 g/kg body weight). Blood samples were drawn from the tail vein and analyzed using a Contour Glucometer (Bayer Pharma AG).

### Cell studies

#### 1. Isolation and culture of primary mouse hepatocytes

Primary mouse hepatocytes were isolated and purified from C57BL/6 mice with a modified two-step perfusion method described in detail previously [32, 33]. After mice anaesthetized, the hepatic portal vein was cannulated and perfused with Kreb’s Ringer containing collagenase IV for 10 min. After the first wash, a second Kreb’s Ringer wash containing CaCl2 and Liberase™ was used for 10 minutes. All solutions were warmed to 37°C. Hepatocytes were filtered through a gauze mesh and resuspended in DMEM (Dulbecco’s modified Eagle’s medium), containing 10% fetal bovine serum, 1 U/mL of penicillin and 1 mg/mL of streptomycin. Cells were plated at six orifice in an atmosphere containing 5% CO2 at 37 °C.

#### 2. Culture of HepG2 cells and intervention for cells

The human hepatoma cell line (HepG2) was cultured in DMEM at an atmosphere containing 5% CO2 at 37 °C. HepG2 cells were passaged every 3 days. Both of the primary mouse hepatocytes and HepG2 cells were divided into olanzapine group and control group. In the olanzapine group, 25, 50, 100 μmol/L olanzapine were respectively added into the medium for intervention for 24 hours. Olanzapine was dissolved in pure DMSO (D2650, sigma, USA). Therefore, in the vehicle, only 0.1% DMSO was added.

### Western blot and QPCR analysis

#### 1. Western blot analysis

For the cell line samples, the cells were washed with ice-cold PBS for three times. RIPA lysis buffer (P0013D, Beyotime Biotechnology, China) containing 1% 0.5 mM phenylmethanesulfonyl fluoride (PMSF) were added, and the cells were incubated at 4 °C for 30 min. The cells were collected and centrifuged at 13,000 g for 15 min at 4°C after been mixed with a pipette. The liver tissues were homogenated in RIPA lysis buffer (P0013B, Beyotime Biotechnology, China) containing 1% 0.5 mMol/L phenylmethanesulfonyl fluoride was added at 100 mg/mL, and then incubated for 30min. Homogenate was centrifugated at 13000 rpm for 15 min at 4°C, the supernatant was saved as a protein extract, and the protein levels were quantified using the BCA protein assay kit (CW0014S, CWBIO, China). Protein extracts were mixed with loading buffer and phosphate buffer saline (PBS), and then the mixture was incubated at 95°C for 5 min for degeneration. Protein extracts were separated by SDS-PAGE and transferred to a PVDF membrane. After blocking with 5% skimmed milk in TBST with 0.1% Tween 20 for 2h at room temperature, the membranes were incubated with primary antibodies overnight at 4°C. The primary antibodies against apoA5 (#3335, Cell Signaling technology, USA; sc-373950, Santa Cruz, USA) and tubulin (66031-1-lg, Proteintech, USA) were diluted 1:1000 in 5% milk/-TBST. The secondary goat anti-mouse and anti-rabbit antibodies (SA00001-2, Proteintech, USA; SA00001-1, Proteintech, USA) were diluted 1:5,000 in milk-TBST, and the membranes were incubated with the corresponding secondary antibodies. The bound complexes were detected with Pierce™ ECL Western Blotting Substrate (32209, Thermo Scientific, USA) and quantified by a Molecular Imager ChemiDoc™ XRS+ (Bio-Rad).

#### 2. QPCR analysis

To further explore the effect of olanzapine on hepatic apoA5 metabolism, *APOA5* mRNA expression levels were measured by quantitative real-time PCR (QPCR). Total RNA was extracted from mouse liver tissues, primary mouse hepatocytes and HepG2 cells according to the protocol of the RNA extraction kit (K0731, Thermo Scientific, USA). First-strand cDNA was synthesized with the Revert Aid First Strand cDNA Synthesis Kit (K1622, Thermo Scientific, USA). QPCR was performed with the SYBR Green Select Master Mix (172-5121, Bio-rad, USA). *GAPDH* were used as the endogenous controls. The primer were synthesized by Tsingke Biological Technology. The sequences of sense and antisense primers were shown in Table 1.

**Table1.**
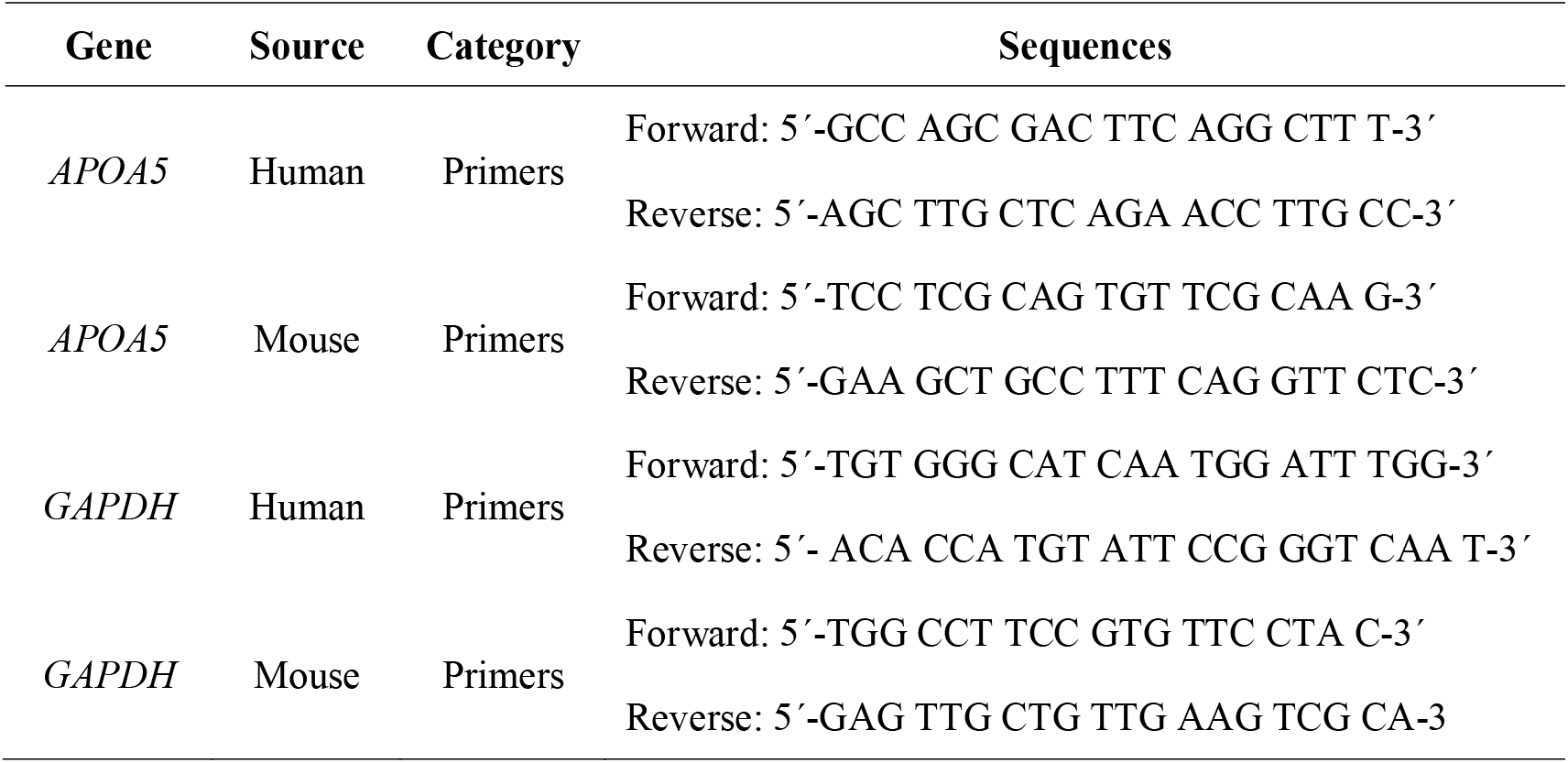
Oligonucleotide sequences of primers and shRNA targeting human and APOA5 gene

### Statistical analysis for animal and in vitro studies

All statistical analyses were carried out with the Graphpad Prism software, and the data was expressed as the mean ± SEM. Statistical analysis was conducted via one-way analysis of variance followed by Dunnett’s t-test, and differences were considered statistically significant at *P* < 0.05.

## Results

### Olanzapine-induced hypertriglyceridemia is caused by decreasing plasma apoA5 concentrations in schizophrenia patients

We characterized the plasma triglyceride and apoA5 levels in 36 drug-naive and first-episode schizophrenic patients treated with olanzapine monotherapy for 2 months. As shown in Figure 1 and Table 2, after olanzapine treatment, plasma triglyceride levels increased and plasma apoA5 levels decreased significantly at each follow-up session. The plasma levels of triglyceride increased as early as 4 weeks. Over the 8-week study period, the mean triglyceride levels increased by 0.76 mmol/l and the mean of apoA5 levels decreased by 48.87 ng/ml. At 8 weeks, 52.78***%*** patients had hypertriglyceridemia (plasma triglycerides ≥ 1.7 mmol/l). To investigate the potential role of apoA5 in olanzapine-induced dyslipidemia, correlation analyses were carried out between plasma apoA5 and lipid levels. It demonstrated that plasma apoA5 levels were significantly negatively correlated with plasma triglyceride levels at 8 weeks (R2 = 0.134, *P* = 0.028) (Figure 1, C) after olanzapine treatment, with no correlation indicated between weight gain and plasma triglycerides (Figure 1, D). On stepwise multiple regression analysis of the independent correlation for triglycerides, plasma apoA5 levels were independently associated with triglycerides at 4 (P = 0.001) and 8 weeks (P = 0.028) after olanzapine treatment (Table 3).

**Table2.** Clinical and biochemical characteristics in schizophrenia patients receiving olanzapine therapy

|  | Baseline | 4 weeks | 8 weeks |
| --- | --- | --- | --- |
| Weight (kg) | 59.72±10.59 | 62.33±11.09 | 65.18±11.23 |
| BMI (kg/m<sup>2</sup>) | 21.39±3.33 | 22.30±3.36 | 23.32±3.33* |
| Systolic blood pressure (mmHg) | 117.61±9.08 | 117.25±9.09 | 116.14±8.69 |
| Diastolic blood pressure (mmHg) | 71.11±6.79 | 71.50±4.79 | 70.78±7.14 |
| **Fasting plasma glucose (mmol/L)** | **4.58±0.69** | **4.56±0.84** | **4.990±0.74** |
| **Triglycerides (mmol/L)** | **1.14±0.60** | **1.51±0.61\*** | **1.90±0.73\*** |
| **Total cholesterol (mmol/L)** | **4.05±0.80** | **4.29±0.71** | **4.49±0.74\*** |
| **HDL cholesterol (mmol/L)** | **1.26±0.34** | **1.29±0.26** | **1.27±0.29** |
| **LDL cholesterol (mmol/L)** | **2.40±0.56** | **2.54±0.53** | **2.71±0.53\*** |
| **Apolipoprotein A5(ng/ml)** | **299.75±48.58** | **265.94±40.51\*** | **250.88±37.35\*** |

\* compared with baseline, $P<0.05$ , body mass index (BMI), high-density lipoprotein cholesterol (HDL), low-density lipoprotein cholesterol (LDL)

**Table3.** Stepwise multiple regression analysis detecting independent contributor to triglyceride in schizophrenia patients

| **Factor** | **$\beta_{\#}$** | **P value** | **$\beta^*$** | **P value** |
| --- | --- | --- | --- | --- |
| **Weight** | **-0.039** | **0.796** | **0.068** | **0.697** |
| **BMI** | **-0.052** | **0.731** | **-0.014** | **0.933** |
| **Fasting plasma glucose** | **-0.272** | **0.060** | **0.062** | **0.707** |
| **Total cholesterol** | **-0.215** | **0.158** | **-0.068** | **0.675** |
| **HDL cholesterol** | **-0.241** | **0.110** | **-0.151** | **0.354** |
| **LDL cholesterol** | **-0.291** | **0.062** | **-0.136** | **0.422** |
| *Apolipoprotein A5* | *-0.534* | *0.001* | *-0.366* | *0.028* |

$\beta$ : Standardized regression Coefficients. $\#$ : 4-weeks vs baseline, \*: 4-weeks vs baseline.

**Figure 1.**
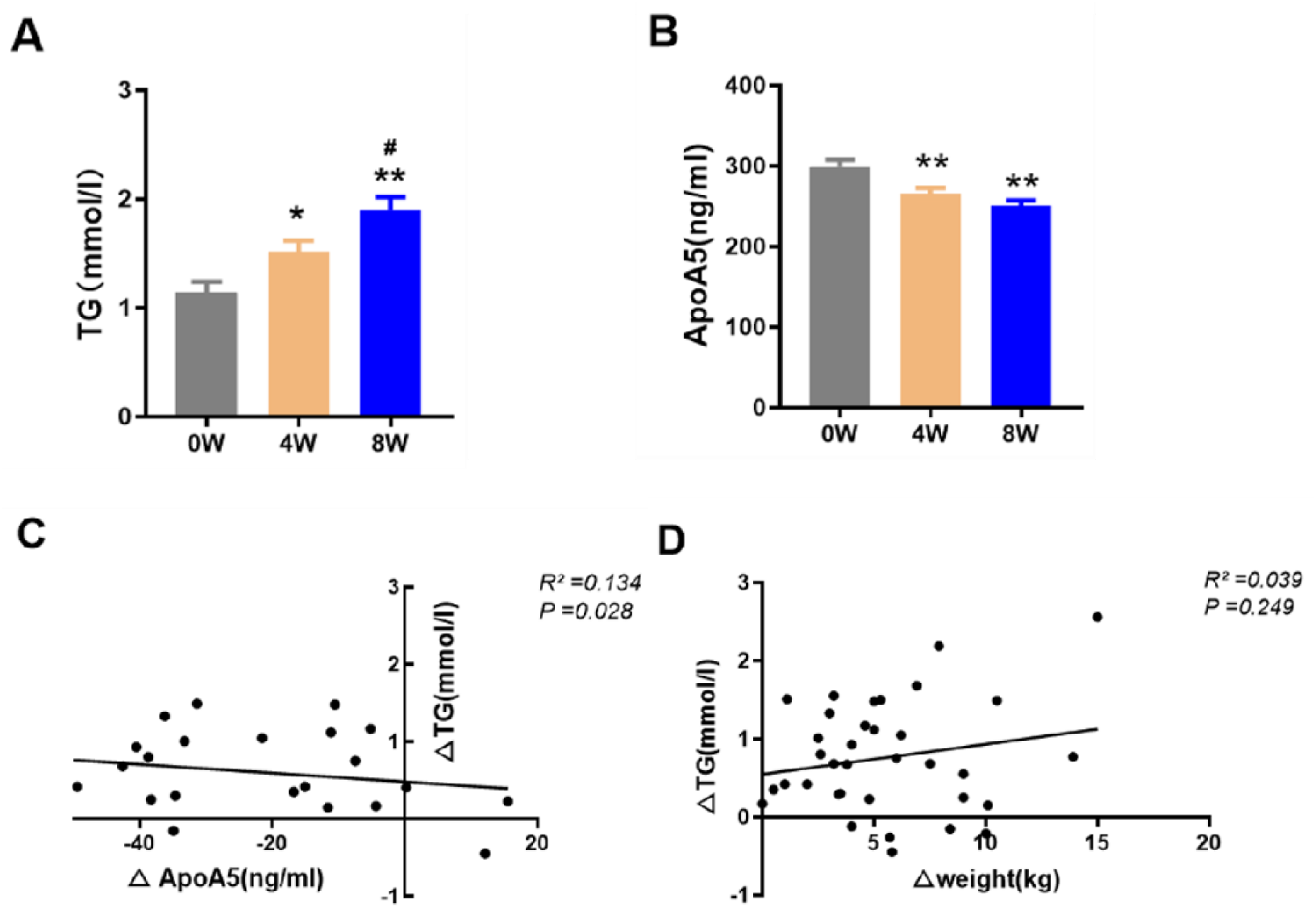
Olanzapine-induced hypertriglyceridemia is caused by decreasing plasma apoA5 levels in schizophrenia patients. (**A**) Triglyceride (TG) levels. **(B)** ApolipoproteinA5 (apoA5) levels. The changes in apoA5 (C) and weight (D) correlated with alterations in triglycerides. Results are shown as mean ± SEM. *P < 0.05 versus control, **P < 0.01 versus control, #P < 0.05 versus 4-weeks olanzapine treatment.

### Changes in body weight and glucose and other lipid outcomes

After 8-week olanzapine treatment, the mean BMI increased significantly by 1.93kg/m2 but all patients’ BMI were within the normal range (18.5 − 24 kg/m2). The plasma levels of total cholesterol, and LDL-C considerably increased, without significant changes in plasma HDL-C and fasting plasma glucose (Supplemental Figure 1, B-E). There is no correlation between the difference between body weight and other metabolic indicators.

#### Effects of olanzapine on body weight, glucose tolerance, blood lipids and apoA5 in mice

Accumulated data have shown that olanzapine treatment leads to weight gain, impaired glucose tolerance and dyslipidemia in mice [34,35]. In this study, we demonstrated that olanzapine treatment contributed to weight gain and impaired glucose tolerance in a dose-dependent manner in mice (Supplemental materials, Figure 2, A -D). Consistent with the observations in humans, increased plasma triglyceride levels were indicated in the two olanzapine groups (199.2 mg/dl in the low-dose olanzapine group and 234.3 mg/dl in the high-dose olanzapine group, versus 131.4 mg/dl in control group, both *P* < 0.05, Figure 2, A). Compared with the low-dose olanzapine group, the elevation of plasma triglyceride levels was more marked in the high-dose olanzapine group (*P* < 0.05). Meanwhile, our results demonstrated that olanzapine treatment dose-dependently decreased plasma apoA5 levels (80.90 ng/ml in the 3 mg/kg olanzapine group, 66.14 ng/ml in the 6 mg/kg olanzapine group, 99.76 ng/ml in control group, all P < 0.05) (Figure 2, B) in animals.

**Figure 2.**
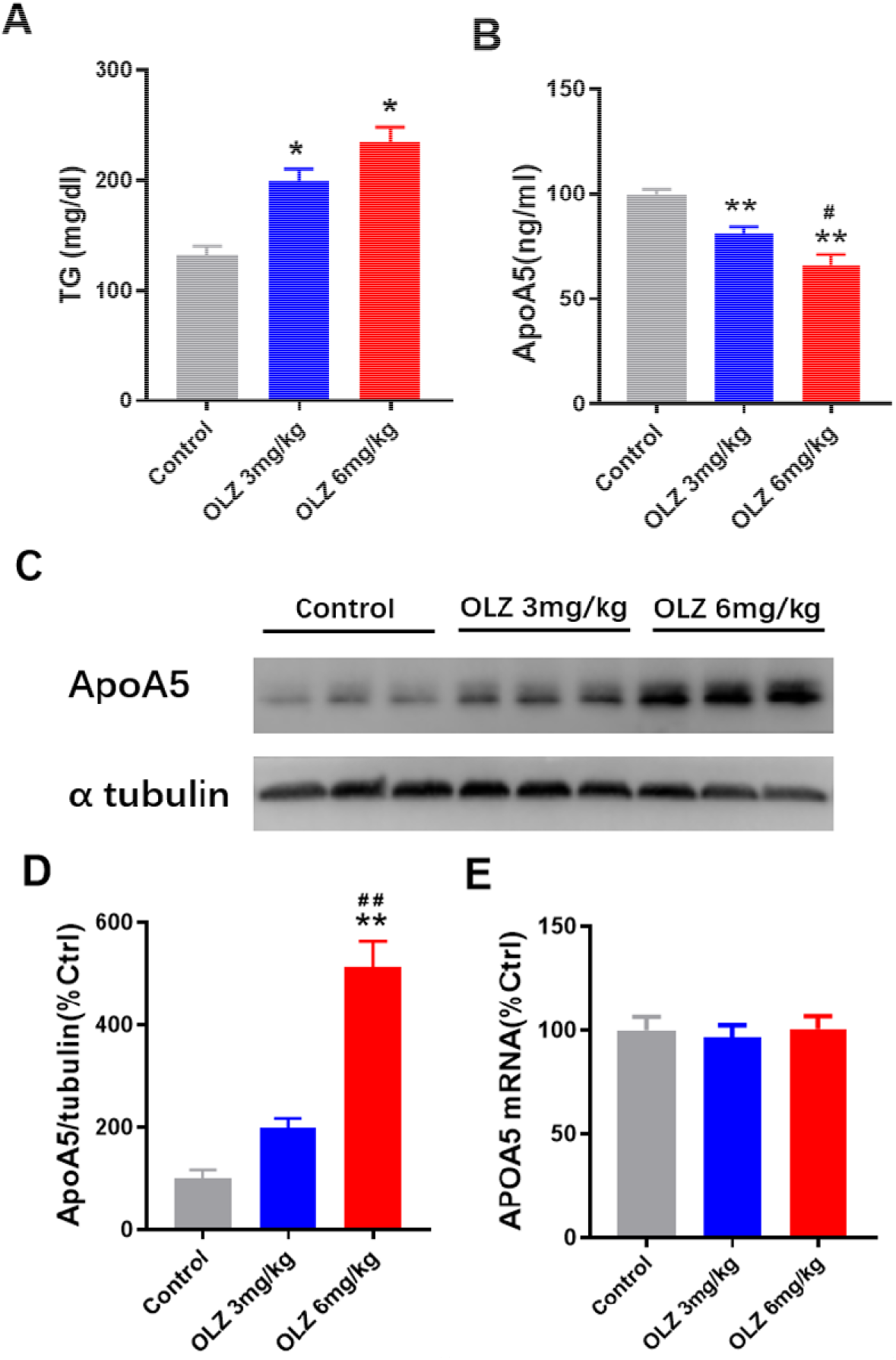
The effect of olanzapine treatment on triglyceride and apoA5 in mice. **(A)** Plasma triglyceride (TG) levels. **(B)** Plasma apoA5 levels by ELISA analysis. **(C and D)** Hepatic apoA5 protein expression in mice by Western blot analysis. **(E)** Hepatic APOA5 mRNA expression in mice by QPCR analysis. Results are shown as mean ± SEM. *P < 0.05 versus control group, **P < 0.01 versus control group, #P < 0.05 versus OLZ 3mg/kg group.

#### Olanzapine increased apoA5 protein levels in mice without changes of APOA5 gene expression

ApoA5 is specifically produced in the liver and hence secreted into bloodstream, in which this protein predominantly plays a triglyceride-lowering role [11, 12]. To investigate the potential role of apoA5 for olanzapine-induced hypertriglyceridemia, we detected apoA5 levels in mice liver. Unpredictably, hepatic apoA5 protein levels dramatically enhanced in mice with 8 weeks of olanzapine treatment (Figure 2, C and D). Interestingly, no statistically significant difference of hepatic *APOA5* mRNA expression was detected among the three groups (*P* > 0.05) (Figure 2, E).

#### Olanzapine increased apoA5 protein levels in hepatocytes in vitro, without changes of hepatocyte APOA5 mRNA levels

To explore the above unpredicted findings of apoA5 in animals, we detected the effects of olanzapine on apoA5 protein and mRNA expression in human and mouse hepatocytes *in vitro*. Similar to the findings in the animal study, our results *in vitro* showed that olanzapine intervention dose-dependently up-regulated apoA5 protein levels in human and mouse hepatocytes cells (Figure 3, A and C). However, we did not found any effect on *APOA5* mRNA levels in hepatocytes *in vitro* (Figure 3, B and D). These observations *in vitro* are in accord with our data *in vivo* from mice, which consistently show that olanzapine administration enhances hepatic apoA5 protein levels instead of hepatic *APOA5* mRNA expression.

**Figure 3.**
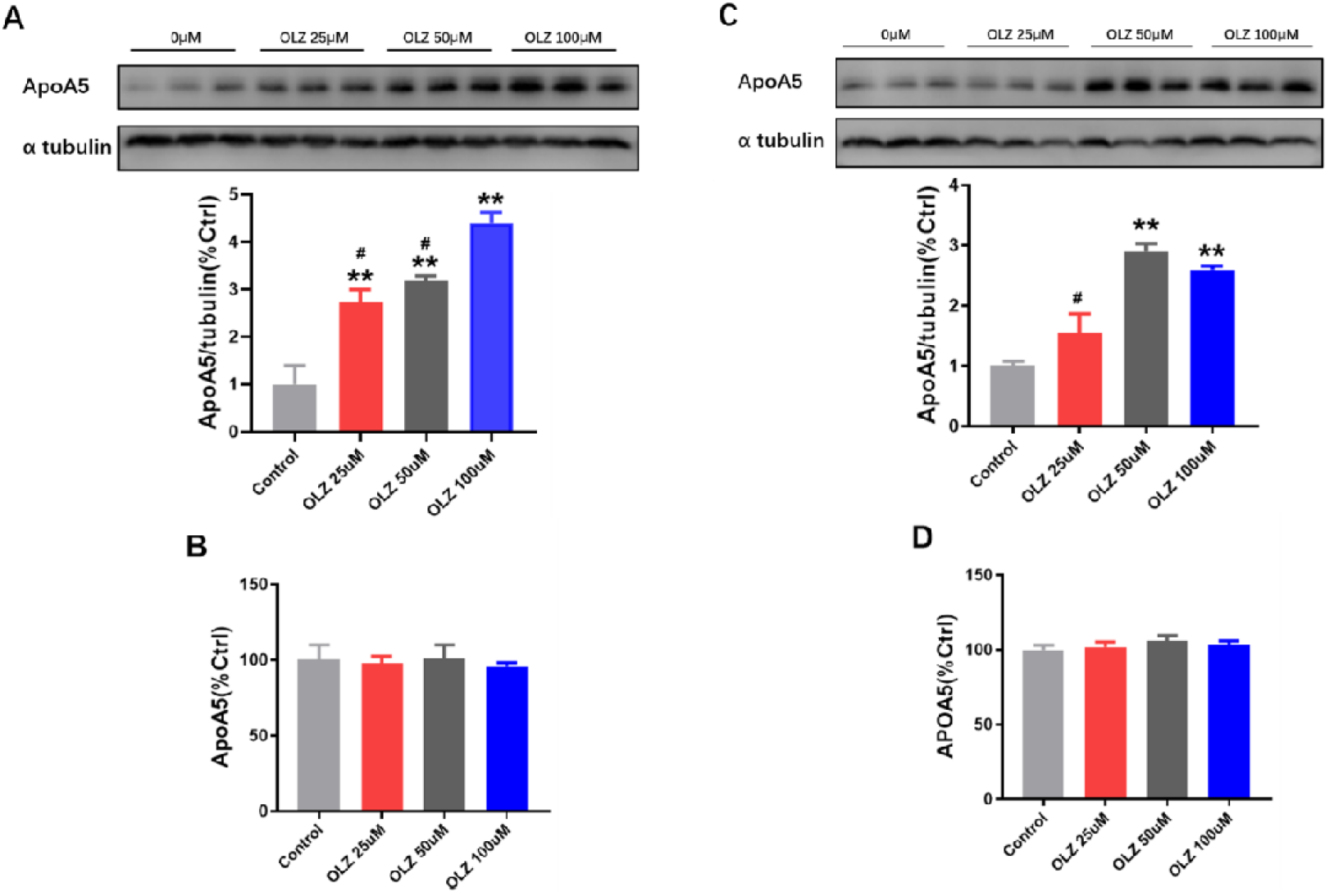
ApoA5 protein and APOA5 mRNA expression in human and mouse hepatocytes in vitro. **(A)** ApoA5 protein expression in HepG2 cells by Western blot analysis. **(B)** APOA5 mRNA expression in HepG2 cells by QPCR analysis. **(C)** ApoA5 protein expression in primary mouse hepatocytes by Western blot analysis. **(D)** APOA5 mRNA expression in primary mouse hepatocytes by QPCR analysis. Results are shown as mean ± SEM. *P < 0.05 versus control group, **P < 0.01 versus control group, #P < 0.05 versus olanzapine (OLZ) 100uM group.

## Discussion

In this study, our findings implicate that inhibition of hepatic apoA5 secretion contributes to olanzapine-induced hypertriglyceridemia. We observed for the first time that olanzapine medication dramatically reduces plasma apoA5 levels which negatively correlated with increased plasma triglyceride levels in schizophrenia patients. Likewise, we got the same finding that olanzapine induced hypertriglyceridemia through decreasing plasma apoA5 levels in mice. Unpredictably, olanzapine remarkably enhanced hepatic apoA5 protein levels and did not change hepatic *APOA5* mRNA expression in mice. Furthermore, similar results of apoA5 protein and mRNA expression were consistently demonstrated in human and mouse hepatocytes in our study *in vitro*. Taken together, our findings suggest that inhibition of hepatic apoA5 secretion (instead of apoA5 production) by olanzapine, resulting in reduced circulating apoA5 levels, leads to hypertriglyceridemia in schizophrenia patients receiving olanzapine medication.

Studies by us and others have previously shown that long-term olanzapine medication predisposes schizophrenia patients to metabolic syndrome (including weight gain, insulin resistance, diabetes and dyslipidemia, *etc*.) that confers a significant increase in risk of CHD [5, 36-38]. Among olanzapine-induced metabolic syndrome, dyslipidemia plays a crucial role in the development of CHD, which typically displays as the proatherogenic dyslipidemia triad, *i*.*e*., elevated plasma triglycerides, decreased HDL-C, and formation of small dense low-density lipoprotein. Among the proatherogenic dyslipidemia triad, hypertriglyceridemia is the key involving the pathogenesis of the other two dyslipidemia, which is acknowledged as an independent risk factor for CHD [39]. Thus, hypertriglyceridemia is regarded as the central olanzapine-induced dyslipidemia. Similarly, our human and animal data in this study indicated that hypertriglyceridemia was the most representative olanzapine-induced dyslipidemia, as evidenced by our previous meta-analysis study [7]. To date, however, the mechanisms underlying olanzapine-induced hypertriglyceridemia remain to be elucidated.

In the present study, apoA5 was enlisted as a candidate contributor for olanzapine-induced hypertriglyceridemia, which is based on the fact that apoA5 has been well-documented as a key regulator for triglyceride metabolism. A large body of animal and human data have implicated apoA5 in reducing plasma triglyceride levels [40-42]. The athero-protective role of apoA5 has also been identified by data from genetically engineered mouse models [16-18], which suggests apoA5 as a potential therapeutic target for treatment of CHD. Specifically, several genetic studies have identified *APOA5* gene polymorphisms as a contributing factor for atypical antipsychotics-associated dyslipidemia [43, 44]. We and others previously also confirmed that up-regulation of hepatic apoA5 expression is responsible for the triglyceride-lowering effect of fibrates [19, 20]. More interestingly, we demonstrated that metformin, a drug for treatment of olanzapine-induced dyslipidemia by our previous data [45], ameliorates obesity-associated hypertriglyceridemia in mice via the apoA5 pathway [46]. Thus, these findings raise the intriguing possibility that apoA5 could participate in olanzapine-related hypertriglyceridemia. In this study, we found that olanzapine treatment considerably increased plasma triglyceride levels and decreased plasma apoA5 levels in schizophrenia patients, with a negative correlation observed between triglycerides and apoA5 levels. Interestingly, however, weight gain was not significantly correlated with plasma triglyceride or apoA5 levels in these patients within the duration of the study, which suggests no marked impact of weight gain on plasma triglyceride or apoA5 levels in this study. It is inconsistent with the previous findings that olanzapine-induced obesity, resulting from diet (high fat diet) and lifestyle (less exercise) changes, participates in dyslipidemia in schizophrenia patients [47]. Notedly, the average BMI of all patients (18.5−24 kg/m2) in our study did not yet reached the criteria of obesity, which is partly due to the short duration of study (*i*.*e*. only 8 weeks of olanzapine treatment). Anyway, our findings indicates an alternative metabolic mechanism for olanzapine-induced hypertriglyceridemia, *i*.*e*., olanzapine, independent of obesity due to diet and lifestyle changes, directly leads to hypertriglyceridemia via the apoA5 pathway.

The triglyceride-lowering effect of apoA5 is mainly involving three pathways. First and the most important, apoA5 is specifically synthesized in the liver and afterward secreted into the bloodstream [11–14], in which apoA5 activates lipoprotein lipase-mediated triglyceride hydrolysis resulting in reduced plasma triglyceride levels [46-48]. Secondly, hepatic apoA5 inhibits the production and secretion of very low-density lipoprotein in the liver [48]. Finally, hepatic apoA5 promotes uptake of triglyceride-rich lipoproteins remnants by the liver, thereby accelerating triglyceride degradation [50]. Given the fact that apoA5 is a liver-specific protein [11, 12], we speculated that reduced plasma apoA5 levels in human and mice in this study could be a consequence of decreased hepatic apoA5 production, which results in olanzapine-induced hypertriglyceridemia. Unexpectedly, however, we found that olanzapine leads to an elevation (not reduction) of apoA5 protein levels in mouse livers and hepatocytes (human and mouse) *in vitro*, but no observed effect of olanzapine on *APOA5* mRNA *in vivo* and *in vitro*. Therefore, our results suggest that reduced plasma apoA5 levels in humans and mice are likely attributed to posttranslational disturbance of hepatic apoA5 protein turnover. Considering the contribution of hepatic apoA5 production and secretion for plasma apoA5 levels, we hold that inhibition of hepatic apoA5 secretion on a posttranslational level, instead of apoA5 production (because of no changes of *APOA5* mRNA expression *in vivo* and *in vitro* by olanzapine), leads to olanzapine-induced hypertriglyceridemia in these schizophrenia patients and mice.

Our above assumption is consistent with the seemingly paradoxical roles of apoA5 in extra- and intra-hepatic triglyceride metabolism, *i*.*e*., apoA5 reduce plasma triglyceride levels and increase hepatocyte triglyceride accumulation [51]. ApoA5 represses hydrolysis of triglycerides and facilitates accumulation of lipid droplets in hepatocytes that promotes the pathogenesis of non-alcoholic fatty liver disease (NAFLD), a disease characterized by excessive triglyceride-rich lipid droplets in hepatocytes [42,52,53]. Pursuant to this, olanzapine significantly attenuates hepatic apoA5 secretion, thereby retaining apoA5 in hepatocytes, which is potentially attributable to the development of NAFLD for schizophrenia patients. Indeed, several lines of data have confirmed that olanzapine induces hepatic lipogenesis that likely increases the susceptibility to NAFLD [54–56]. Concomitantly, olanzapine-induced hepatic lipid accumulation is associated with activation of sterol regulatory element-binding protein [56], a transcription factor involved in hepatic apoA5 expression [57]. Therefore, although it has yet to be determined, these previous data suggest a potential link between olanzapine-induced hepatic apoA5 retention and excess hepatic lipid accumulation that could promote NAFLD pathogenesis in schizophrenia patients.

Here, we propose a hypothesis for the role of apoA5 in olanzapine-induced hypertriglyceridemia (Figure 4). Physiologically, apoA5 protein is synthesized within the endoplasmic reticulum in hepatocytes, transported into the Golgi body, and hence secreted into the blood for reducing circulating triglyceride levels. However, olanzapine significantly attenuates hepatic apoA5 secretion, resulting in reduced plasma apoA5 levels and hence hypertriglyceridemia. Concomitantly, inhibition of hepatic apoA5 secretion will cause hepatocyte apoA5 retention, thereby facilitating biogenesis of triglyceride-rich lipid droplets and therefore leading to the development of NAFLD.

**Figure 4:**
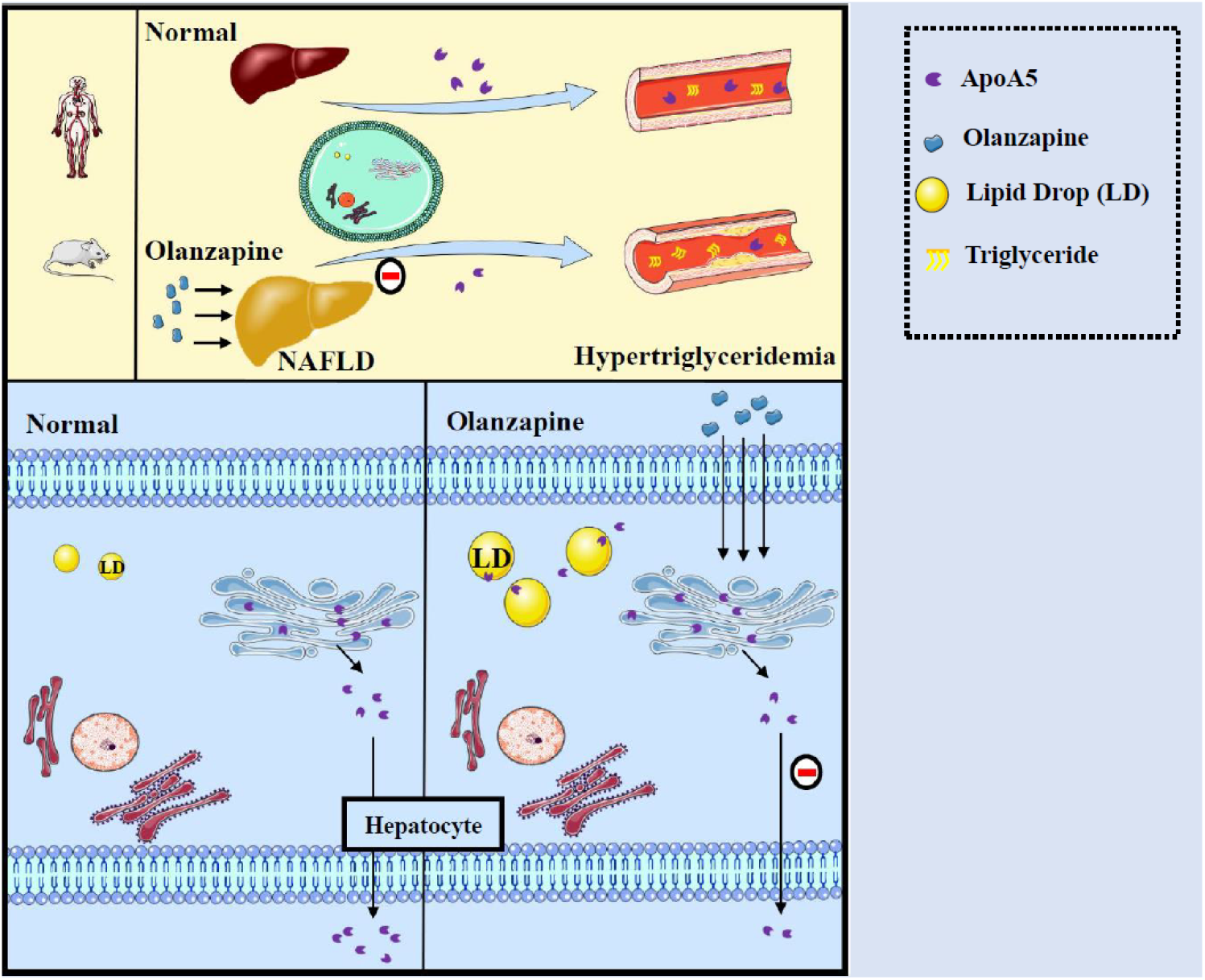
Inhibition of hepatic apoA5 secretion by olanzapine leads to hypertriglyceridemia and potentially NAFLD (non-alcoholic fatty liver disease). ApoA5, apolipoprotein A5; LD, lipid droplet.

## Conclusion

This study for the first time investigated the role of apoA5 in olanzapine-induced hypertriglyceridemia. Our findings indicate that olanzapine treatment inhibits hepatic apoA5 secretion, resulting in decreased plasma apoA5 levels and hence increased plasma triglyceride levels. These data suggests that apoA5 may serve as a target for developing therapeutics for hypertriglyceridemia in schizophrenia patients taking olanzapine medication. However, the mechanisms underlying olanzapine-induced inhibition of hepatic apoA5 secretion remains to be investigated in the future.

## Data Availability

All data used to support the findings of this study are included within the article and supplemental materials.

## Authors and contributors

Xiansheng Huang designed the experiments. Wenqiang Zhu, Piaopiao Huang and Jingmei Xiao performed the experiments. Yang Yang, Li Shen, Fei Luo and Wen Dai analysed the data. Xiansheng Huang, Wenqiang Zhu and Piaopiao Huang prepared the initial draft of the manuscript. Renrong Wu and Rong Li revised the text. All authors approved the final draft of the manuscript.

## Acknowledgments

This work was supported by the Key R&D Program Projects, National Science

Foundation of China (Grant No.2016YFC1306900),the National Natural Science

Foundation of China (Grant No. 81974281 and No. 82072096), the Natural Science Foundation of Hunan Province (No.2017JJ2351; No. 2018JJ3741; No.2020JJ2052), the National Natural Science Foundation for Young Programs of China (No. 81700999).

