## Supplemental materials for "Olanzapine inhibits hepatic apolipoprotein A5 secretion inducing hypertriglyceridemia in schizophrenia patients and mice"

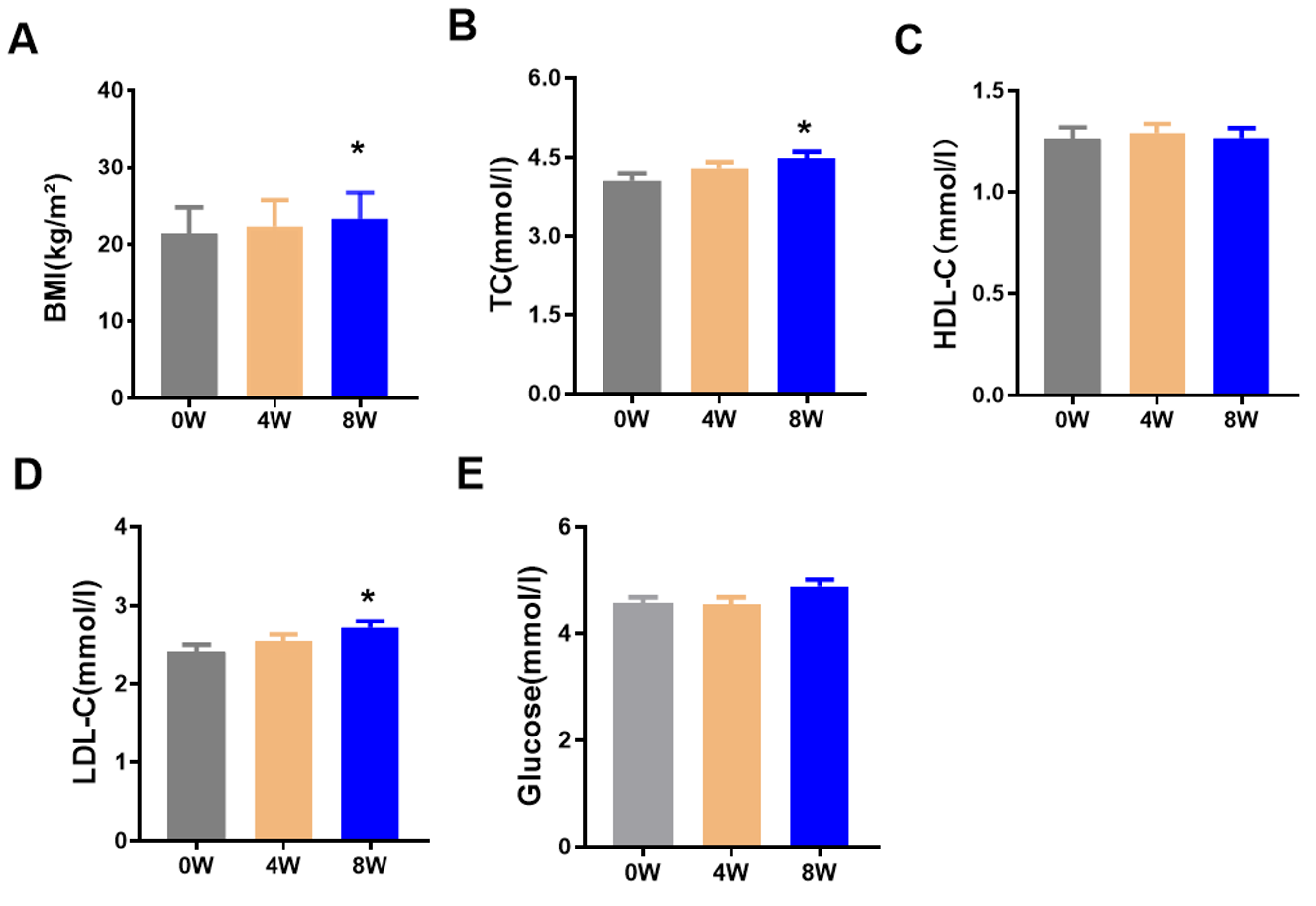

***Figure 1. Effects of olanzapine treatment on plasma lipid and glucose levels in schizophrenia patients.*** *(****A****) BMI (Body mass index). (B) Total cholesterol (TC) levels.* ***(C)*** *High-density lipoprotein cholesterol (HDL-C) levels* ***(D)*** *Low-density lipoprotein cholesterol (LDL-C) levels.* ***(E)*** *Glucose levels. Results are shown as mean ± SEM. *P < 0.05 versus control, **P < 0.01 versus control, ^#^P < 0.05 versus 4-weeks olanzapine treatment.*

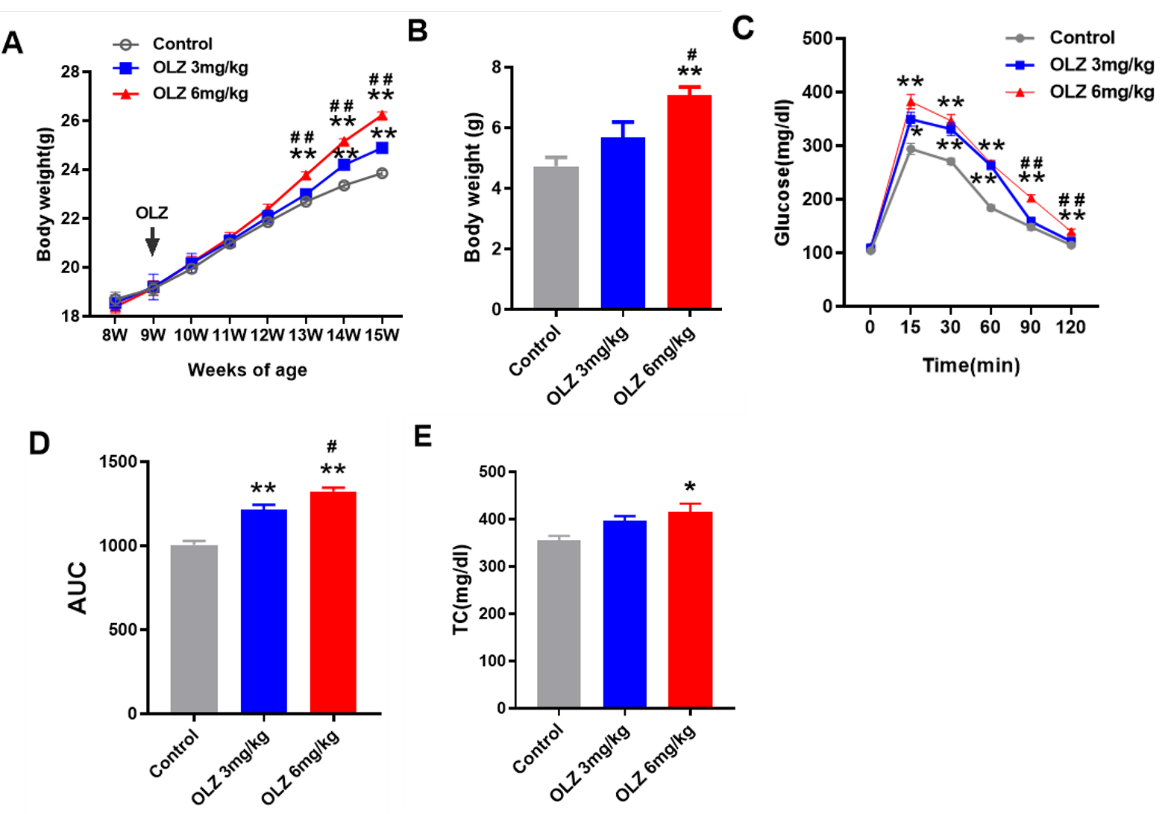

***Figure 2. Effect of olanzapine on body weight, glucose tolerance and blood lipids in mice*** *(n = 7 each group).* ***(A)*** *Body weight.* ***(B)*** *Weight gain from baseline.* ***(C)****Glucose tolerance test (GTT).* ***(D)*** *Area under the curve (AUC) of GTT.* ***(E)*** *Plasma total cholesterol (TC) levels. Results are shown as mean ± SEM. *P < 0.05 versus control group, **P < 0.01 versus control group, # P < 0.05 versus olanzapine (OLZ) 3mg/kg group, ## P < 0.01 versus OLZ 3mg/kg group.*
